# Comments on “EpiRank: Modeling Bidirectional Disease Spread in Asymmetric Commuting Networks” for Analyzing Emerging Coronavirus Epidemic Patterns

**DOI:** 10.1101/2020.04.20.20072736

**Authors:** Wei Chien Benny Chin, Chung-Yuan Huang

## Abstract

Huang et al.^1^ used their EpiRank algorithm, which emphasizes forward-and-backward commuter flow between homes and workplaces, to analyze the distribution patterns of two infectious diseases in Taiwan: the 2009-H1N1 influenza virus and the widespread emergence of the 2000-2008 type 71 enterovirus (EV). As this article was being prepared, the spreading mechanism of the novel coronavirus disease now designated as COVID-19 had yet to be identified, but according to the American Centers for Disease Control, its spreading mechanism and patterns are likely more similar to influenza than to other coronaviruses such as Severe Acute Respiratory Syndrome (SARS-CoV-1) or Middle East Respiratory Syndrome (MERS-CoV). To consider potential COVID-19 spatial patterns, we applied EpiRank to the 2003 SARS outbreak in north Taiwan for comparison with H1N1 and EV. SARS was found to be less contagious than H1N1 or EV, but with a significantly higher fatality rate. The characteristics of these diseases determined their specific spatial spreading patterns, as reflected in the different effects of forward and backward commuting movement. Our motivation is to highlight these differences and to illustrate EpiRank spatial patterns for the 2003 SARS outbreak for comparison with EpiRank-determined distributions for the H1N1 and EV outbreaks. Our results indicate that the daytime parameter (i.e., forward movement effect) range was slightly higher (0.5-0.55) for the SARS outbreak than for either the influenza (0.4-0.5) or EV (0.3-0.5) outbreaks, suggesting that the forward-and-backward movements of individuals between residential and core urban areas with concentrated populations were equally important regarding the spread of SARS. While COVID-19 might resemble either SARS or H1N1 in terms of spatial spreading, its daytime parameter is likely somewhere in-between, with backward movement being dominant (similar to H1N1) or with forward and backward movement being equally important (similar to SARS). Building on Huang et al.’s paper, we present an estimated risk distribution pattern for the Taipei Metropolitan Area for a daytime parameter of 0.55.

## Introduction

The virological characteristics of Coronavirus Disease 2019 (COVID-19) have high degrees of similarity with the 2003 Severe Acute Respiratory Syndrome Coronavirus (SARS-CoV-1, 79% identity) and 2012 Middle East Respiratory Syndrome Coronavirus (MERS-CoV, approximately 50% identity)^2^. However, in mid-February 2020 the COVID-19 spreading mechanism and spatial epidemiology were both believed to be more similar to influenza and other respiratory infections in terms of primary transmission method—respiratory droplets^3^. Infected patients can be asymptomatic or express a broad range (mild to severe) of COVID-19 symptoms^4^. Similar to influenza, COVID-19 carriers can transmit the disease whether or not they show symptoms—a characteristic of special concern to epidemiologists, public health specialists, and government officials^5,6^. The worst-case scenario is that this asymptomatic transmission characteristic can result in an international COVID-19 pandemic that would be as difficult to control as the 2003 SARS outbreak and prior influenza pandemics^7^.

Huang et al.^1^ created the EpiRank algorithm to investigate the spreading of diseases via inter-city commuting networks. EpiRank integrates the effects of transmission via the forward and backward movements of commuters. Its primary assumption is that initial carriers move the virus from homes to workplaces (forward), infect other commuters using public transportation as well as coworkers and susceptible individuals during their daily activities, and then infect others during the return commute and family members in their homes. EpiRank calculations use a “daytime parameter” to capture the effect of forward and backward movement, and a “damping factor” to capture diffusion proportions according to commuting network characteristics rather than random transmission. Their study focused on the 2009 H1N1 (Influenza A) outbreak and the spatial distribution of the 2000-2008 type 71 enterovirus (EV), both in Taiwan. Their results revealed optimum daytime values between 0.3 (Pearson’s R) and 0.5 (Spearman’s rho) for flu cases and between 0.4 (Pearson’s R) and 0.5 (Spearman’s rho) for EV cases. Daytime values below 0.5 were viewed as evidence of a stronger backward movement effect on the spreading of flu and EV cases.

Our motivation for this paper is to expand Huang et al.’s EpiRank analysis^1^ to the 2003 SARS outbreak in Taipei in order to offer a reference for other SARS-like diseases. COVID-19 is similar to SARS in terms of virology, and although it may be more similar to influenza in terms of its spatial spreading mechanism, we believe that providing EpiRank results for SARS, in addition to the flu and EV cases covered in Huang et al.’s original paper, can support future COVID-19 intervention decisions.

### 2003 SARS outbreak in Taipei

In our earlier study we used data for the Taipei Metropolitan Area (TMA) to test the sensitivity of the EpiRank daytime parameter and damping factor when assessing risk distribution associated with commuting flow. Our intention in this paper is to provide additional insights into spatial pattern differences among the 2003 SARS outbreak, 2009 H1N1 outbreak, and EV cases recorded between 2000 and 2008.

A total of 347 SARS cases were confirmed in Taiwan in 2003, with 81% (282) occurring in three urban centers in the northern part of the country: Taipei, New Taipei City, and Keelung^8^. For the present analysis we focused on the 48 townships located within or surrounding those three centers. In this section we will describe our sensitivity analysis involving the daytime parameter and damping factor for investigating the effects of morning and evening commutes on the spread of SARS, and then discuss the effects of spreading via commuting networks as opposed to random infectivity. We will then draw and explain two SARS/EpiRank distribution maps reflecting optimized parameters.

The sensitivity analysis results shown in Figs. 1a and b respectively present Pearson’s and Spearman correlation data for the actual distribution of SARS cases and EpiRank-predicted distributions based on different ranges of values for damping factors (X-axis) and daytime parameters (Y-axis). They reveal significant correlations between the two when the daytime parameter was set to 0.5 or 0.55 and the damping factor set to 1. As in Huang et al.’s experiments^1^, we only considered a uniform probability distribution for the external factor—that is, all nodes had the same probability of coming into contact with a contagious individual. Accordingly, a damping factor of 1 means that disease transmission was more dependent on network structure than a uniform distribution. A daytime parameter approaching 0.55 indicates that the effects of both forward and backward movements were essentially equal in terms of disease diffusion, with forward movement only slightly more important.

**Figure 1.**
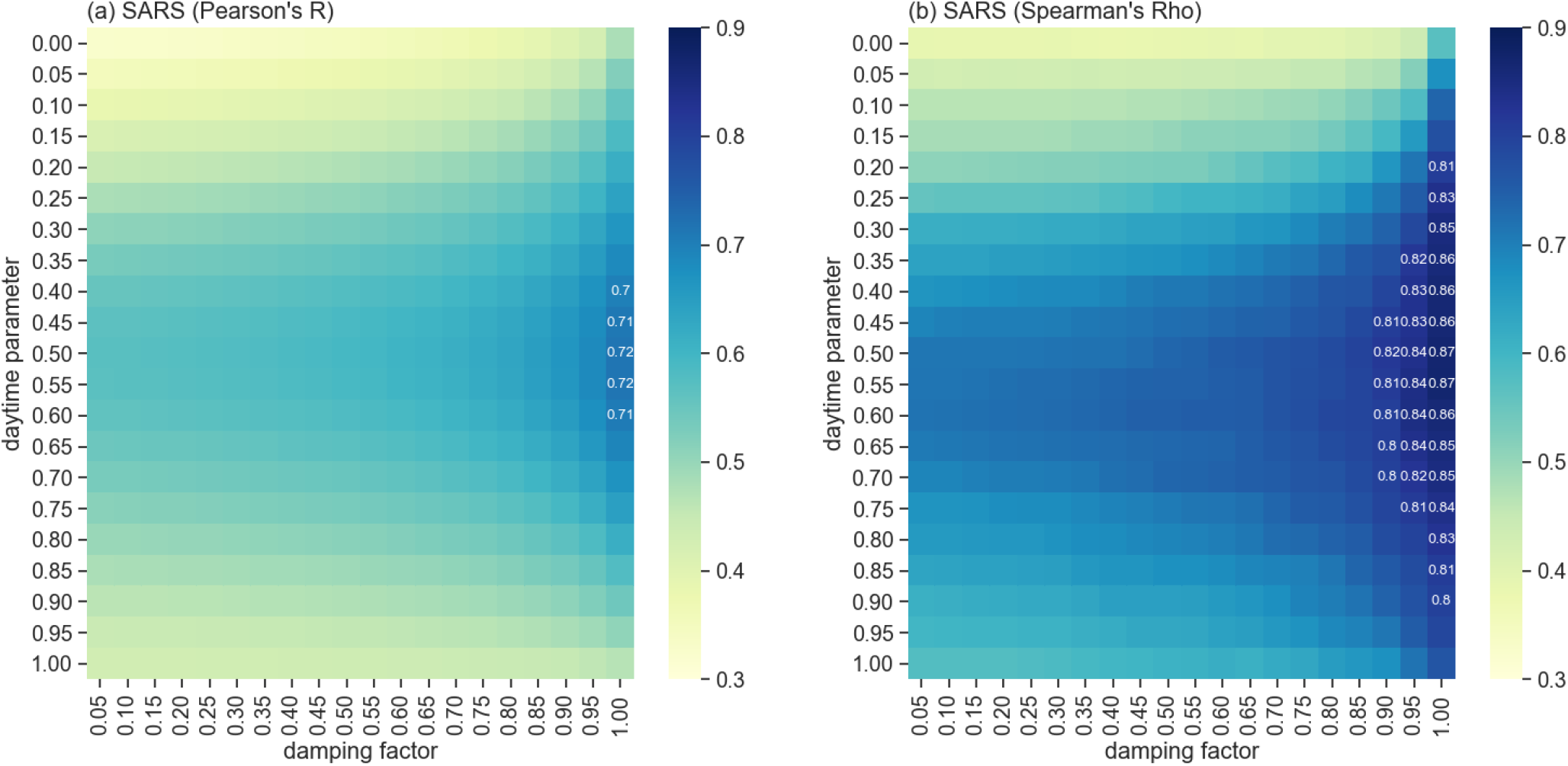
Sensitivity data for daytime (forward and backward movement) and damping factor values (effect of network topology compared to external factors). (a) Pearson’s R, (b) Spearman’s rho.

The structure of the SARS sensitivity pattern shown in Fig. 1 is the same used by Huang et al.^1^ for the flu and EV daytime parameters in Fig. 2—that is, optimized daytime parameters for flu of 0.4-0.5 and for EV of 0.3-0.5. All of these values are at or below 0.5, indicating a stronger backward movement effect. Possible explanations for the higher SARS daytime parameter (>0.5) that we observed in the present study include a higher fatality rate, smaller number of patients, effects of public health policies, and a slightly lower basic reproduction number (*R*_0_). In Taiwan, SARS had a much higher fatality rate than flu (13%^9^ versus 0.001%^10^), and therefore attracted greater media and research attention over a shorter time period. The fatality rate triggered a rapid government response in the form of several policy decisions involving large amounts of resources aimed at controlling the disease. These policies resulted in a much lower overall number of SARS patients compared to flu patients.

**Figure 2.**
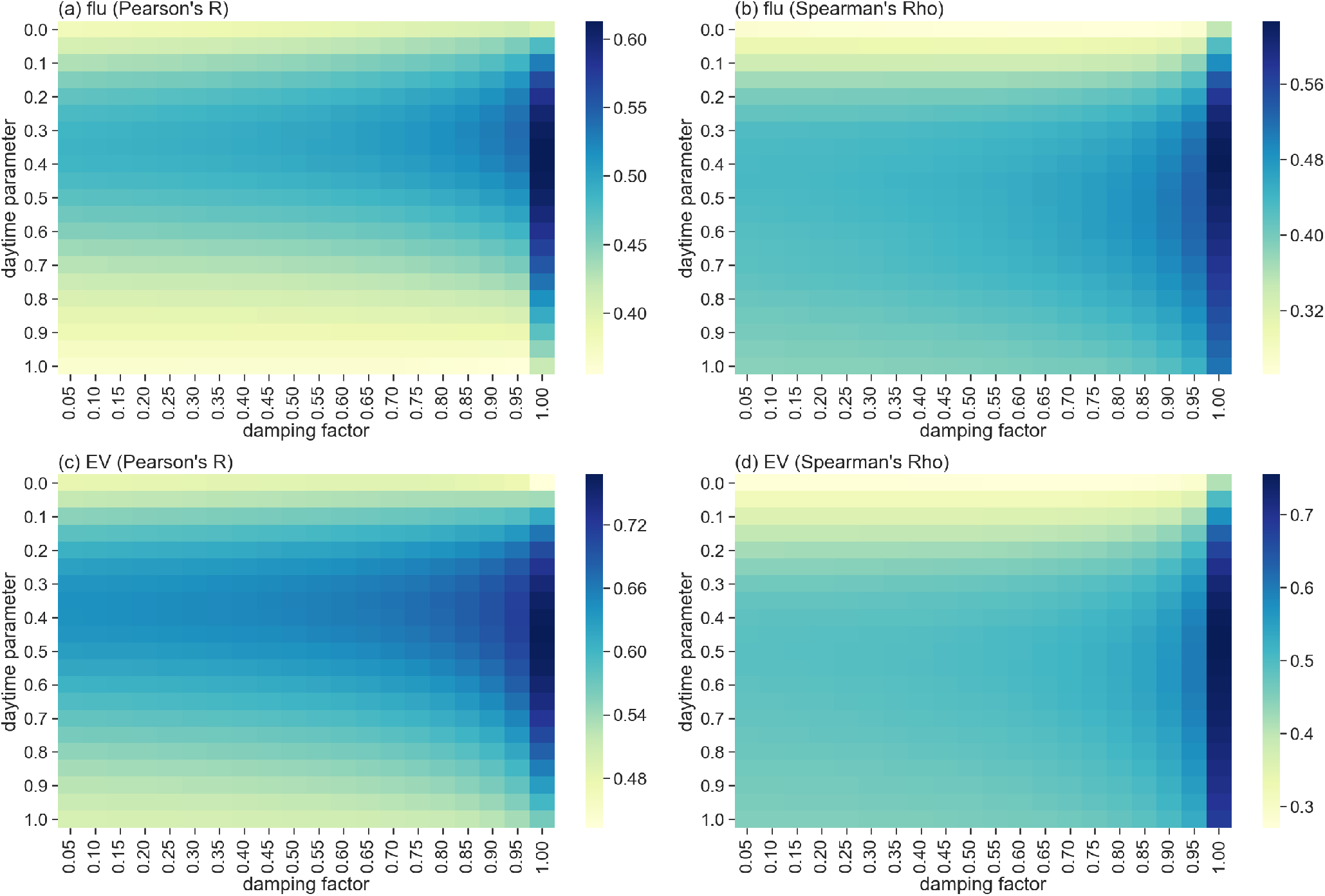
Sensitivity data for daytime (forward and backward movement) and damping factor values (effect of network topology compared to external factors) with the flu and EV cases. This figure was replicated from Huang et al.^1^. (a) Pearson’s R correlation with flu, (b) Spearman’s rho correlation with flu. (c) Pearson’s R correlation with EV, (b) Spearman’s rho correlation with EV.

*R*_0_, defined as the expected number of cases caused by a single infected patient, is used to describe the spreading capabilities of diseases. The *R*_0_ values associated with the H1N1 virus in Taiwan were 1.16 during the first phase and 1.87 during the second^11^; the SARS *R*_0_ was measured as 1.54^12^—in other words, the 2009 H1N1 influenza strain was more contagious than SARS. Patients infected with H1N1 could carry disease pathogens with few or no discernable symptoms, increasing the potential to infect susceptible individuals—especially coworkers and family members. However, when they showed signs of the disease, they were more likely to stay at home and treat themselves. During the SARS outbreak, individuals who exhibited even the faintest symptoms were immediately sent to hospitals for treatment, which mitigated the spreading potential. This difference explains, at least in part, why the backward movement was more important for influenza than for SARS.

### 2003-SARS spatial distribution identified by EpiRank

We used head/tail-breaks^13^ to separate the 48 northern Taiwan townships into four core-levels: I, II, III and non-core. Core levels associated with the actual SARS and EpiRank-predicted spatial distributions are shown in Fig. 3. SARS cases were mostly concentrated in 1 core-I township and 5 core-II townships near or just outside of southwest Taipei (Fig 3a). The core-I township (Wanhua district) had 44 cases (15.6% of all cases in the TMA), and each of the five core-II townships (2 in Taipei, 3 in New Taipei City) had between 21 and 26 cases (7.4%-9.2%). We then fed distribution data into an optimized EpiRank with the daytime parameter set to 0.55 and damping factor to 1.0 (Fig. 3b). The EpiRank results identified 5 core-I townships (5.2%-6.9% of total EpiRank score), 4 core-II townships, and 1 core-III township later identified as the Wanhua district (2.8%).

**Figure 3.**
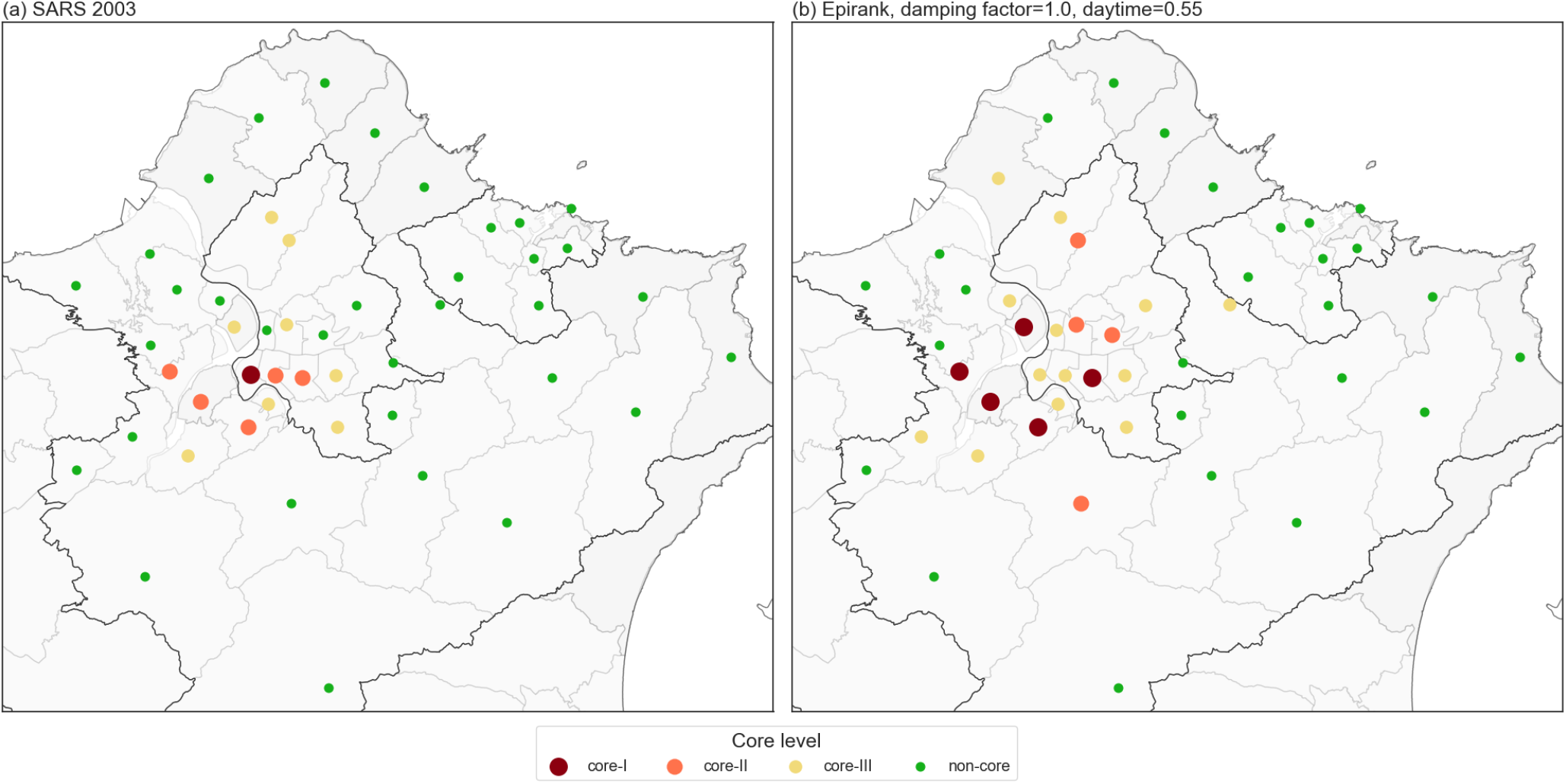
(a) Actual core-I, core-II, core-III and non-core township locations for the 2003 SARS outbreak. (b) Core-I, core-II, core-III and non-core townships identified by EpiRank.

Wanhua, the epicenter of the 2003 SARS outbreak in Taiwan^14^, had approximately 1.5 times more cases than the second highest township. Looking at the north Taiwan commuting network, neither human activity nor population density levels in Wanhua were as high as in surrounding EpiRank-identified core-II townships, namely the Daan district of Taipei and the Banqiao, Sanchong, Zhonghe and Xinzhuang districts of New Taipei City. These five townships are major TMA residential centers with large numbers of daily commuters to the Taipei core. Accordingly, they were identified as having the highest SARS infection risks based on results from a commuting network analysis.

## Summary

EpiRank was designed to serve as a useful tool for analyzing commuting flows—that is, the daily forward and backward movements of individuals within regions. Since the COVID-19 spreading mechanism and spatial diffusion process have yet to be firmly identified, we returned to our earlier analysis of three events involving infective diseases in Taiwan: the 2003 SARS outbreak, the 2009 H1N1 influenza outbreak, and the large number of type 71 enterovirus (EV) cases between 2000-2008. H1N1 and EV were part of Huang et al.’s original EpiRank research^1^. Our results indicate daytime parameter values (i.e., measures of the home-to-work movement effect on disease spreading) of between 0.5 and 0.55 for the SARS outbreak, higher than those observed for the flu and EV scenarios (both <0.5). Even though SARS and influenza are both respiratory illnesses, they are distinctly different in terms of fatality rates and contagiousness, with the second factor affecting the variation we observed in disease spreading patterns. Since the COVID-19 epidemic is still emerging (with rapidly evolving information regarding mortality and contagiousness), it is not possible to accurately determine the effect of commuting on disease spread—that is, whether it is similar to SARS or flu. For this project we applied EpiRank to the SARS case for comparison with Huang et al.’s earlier work with influenza^1^.

Some of the earliest analyses of COVID-19 suggest that it is more similar to seasonal influenza than SARS in terms of its spatial spreading mechanism. If true, the backward movement of individuals living in and close to the epicenter of the COVID-19 outbreak will likely exert more effects than their forward movement, with greater likelihood of COVID-19 spreading from workplaces and other city center locations to residential neighborhoods. In the present study we observed equal effects of forward and backward movement (daytime parameter = 0.55) on the spatial distribution of estimated disease risk, which resembles the situation for the 2003 SARS outbreak in Taiwan. In our original study we discussed an estimated risk distribution with the largest possible backward movement effect (daytime parameter = 0.0).

Since EpiRank uses commuting network flow data, it may be useful for identifying disease spreading patterns and high at-risk communities even when data are lacking for other epidemiological factors or important variables. As more data become available for commuting flows in China in general and Wuhan in particular, EpiRank may have utility for analyzing actual and potential disease diffusion, as well as for creating estimated risk distribution maps for early response and control purposes. EpiRank might also be applied to other population flow networks for analyzing risk distribution involving routine patterns (e.g., global flight networks). Since it was written using Python 3, EpiRank can easily be applied to other scenarios and locations where commuting network flow data are available.

In our role as researchers specializing in network-based epidemiologic modelling and simulations, we encourage the public to fully support all government-sponsored responses to and preventive measures against COVID-19. As of this writing the outbreak has not yet resulted in any infection clusters in Taiwan, therefore now is the time to organize and prioritize disease prevention and medical treatment resources for frontline medical personnel. EpiRank can serve as a reference tool for allocating limited resources to potential areas of infection in case a COVID-19 outbreak does occur—that is, EpiRank can be used to determine where resources should be concentrated based on epidemiologic and economic priorities. The guiding goal is to fully contain a COVID-19 outbreak in Taiwan at the earliest possible stage.

## Data Availability Statement

All processed data and algorithm codes are available at https://github.com/wcchin/EpiRank.

## Data Availability

All processed data and algorithm codes are available at https://github.com/wcchin/EpiRank.

https://epirank-app.herokuapp.com/

## Acknowledgements

This work was supported by the Republic of China Ministry of Science and Technology (MOST-107-2221-E-182-069) and the High Speed Intelligent Communication (HSIC) Research Center of Chang Gung University, Taiwan.

## Author contributions statement

C.Y.H. organized the data, W.C.B.C. ran the analysis and organized the results, and W.C.B.C. and C.Y.H. wrote the manuscript.

## Additional information

### Competing Interests

The authors declare no competing financial interests.

## Notes

### Competing Interest Statement

The authors have declared no competing interest.

